# Evidence of the effectiveness of travel-related measures during the early phase of the COVID- 19 pandemic: a rapid systematic review

**DOI:** 10.1101/2020.11.23.20236703

**Authors:** Karen A. Grépin, Tsi Lok Ho, Zhihan Liu, Summer Marion, Julianne Piper, Catherine Z. Worsnop, Kelley Lee

## Abstract

**Objective:** To review evidence of the effectiveness of travel measures implemented during the early stages of the COVID-19 pandemic in order to recommend change on how evidence is incorporated in the International Health Regulations (2005) (IHR).

**Design:** We used an abbreviated preferred reporting items for systematic reviews and meta-analysis protocol (PRISMA-P) and a search strategy aimed to identify studies that investigated the effectiveness of travel-related measures (advice, entry and exit screening, medical examination or vaccination requirements, isolation or quarantine, the refusal of entry, and entry restrictions), pre-printed or published by June 1, 2020.

**Results:** We identified 29 studies, of which 26 were modelled (vs. observational). Thirteen studies investigated international measures while 17 investigated domestic measures (one investigated both), including suspended transportation (24 studies), border restrictions (21), and screening (5). There was a high level of agreement that the adoption of travel measures led to important changes in the dynamics of the early phases of the COVID-19 pandemic. However, most of the identified studies investigated the initial export of cases out of Wuhan, which was found to be highly effective, but few studies investigated the effectiveness of measures implemented in other contexts. Early implementation was identified as a determinant of effectiveness. Most studies of international travel measures failed to account for domestic travel measures, and thus likely led to biased estimates. Poor data and other factors contributed to the low quality of the studies identified.

**Conclusion:** Travel measures, especially those implemented in Wuhan, played a key role in shaping the early transmission dynamics of the COVID-19 pandemic, however, the effectiveness of these measures was short-lived. There is an urgent need to address important evidence gaps, but also a need to review the way in which evidence is incorporated in the IHR in the early phases of a novel infectious disease outbreak.

**What is already known on this subject?:**

- Previous reviews of the evidence from outbreaks of influenza and other infectious disease have generally found that there is limited evidence that travel-measures are effective at containing outbreaks.
- However, it is unclear if the lessons from other infectious disease outbreaks would be relevant in the context of COVID-19.
- Based on evidence at the time, WHO did not recommend any travel restrictions when it declared COVID-19 a Public Health Emergency of International Concern.

**What does this study add?:**

- This study rapidly reviews the evidence on the effectiveness of travel measures implemented in the early phase of the pandemic on epidemiological countries.
- The study investigated both international and domestic travel measures and a wide range of travel measures.
- The study finds that the domestic travel measures implemented in Wuhan were effective at reducing the importation of cases internationally and within China. The study also finds that travel measures are more effective when implemented earlier in the outbreak.
- The findings generate recommendations on how to incorporate evidence into the International Health Regulations and highlights important research gaps that remain.

**How might this affect future outbreaks?:**

- The findings of this study suggest the need to decouple recommendations of travel measures from the declaration of a public health emergency of international concern.
- Highlights the need to evaluate the potential effectiveness of travel measures for each outbreak, and not just assume effectiveness based on past outbreak scnearios.

## Introduction

On January 31, 2020, the World Health Organization (WHO) declared the outbreak of SARS-CoV-2 a Public Health Emergency of International Concern (PHEIC). Based on information available at the time, the International Health Regulations 2005 (IHR) Emergency Committee explicitly did not recommend “any travel or trade restriction”.^1^ The IHR state that State Parties should avoid unnecessary interference with international traffic in the adoption of measures and that the measures should not be “more restrictive of international traffic and not more invasive or intrusive to persons than reasonably available alternatives that would achieve the appropriate level of health protection” (Article 43). Moreover, measures adopted should be based on “scientific principles,” evidence and/or WHO guidance.

As early as December 31, 2019, the same day that China Centre for Disease Control first notified WHO of a cluster of atypical pneumonia cases in Wuhan, some authorities (including Taiwan, Russia, and Macau) began to impose targeted travel-related measures, mainly airport screening of travellers from Wuhan.^2^ Within weeks, additional locations also restricted flights to or suspended entry from Wuhan, including Mongolia, Australia, and North Korea. On January 23, a *cordon sanitaire* was drawn around Wuhan, effectively suspending all international and domestic travel in and out of the city.^3^ A day later, the measures were extended to all of Hubei province. By March 2020, despite WHO’s recommendations, virtually all IHR (2005) States Parties had implemented some form of cross-border travel-related measure in response to the COVID-19 pandemic.^4^ This is, by far, the largest number of countries adopting such measures during a PHEIC: only about a quarter of countries had imposed such measures during the 2009 H1N1 pandemic and the 2014 Ebola outbreak in West Africa.^5–7^ It is estimated that there was a 65% drop in international travel in the first half of 2020 as a result of the COVID-19 pandemic.^8^

The near universal adoption of travel-related measures, especially in light of the potentially large economic and social consequences, raises questions as to whether such measures can be, and have been, effective at reducing international transmission of the virus during the pandemic. Previous studies have suggested that certain travel-related measures have only limited, or at best modest, effectiveness in containing outbreaks of influenza. A systematic review of the effectiveness of international travel measures (screening, travel restrictions, and border closures) to control pandemic influenza identified only 15 studies and found that measures implemented early could delay local transmission by a few days or weeks, slow international spread, and delay the epidemic peak in isolated locations by reducing the number of seeding events.^9^ The review did not identify any evidence that screening methods were effective but it did find that border closures had been effective in preventing virus introduction to small island states during the 1918 influenza pandemic. However, the authors also concluded that the overall evidence base on which they drew their conclusions was small and of low quality.

A related but larger review of a broader range of measures, including travel advice, screening, internal travel restrictions, and border closures, for both epidemic and pandemic influenza, also found that travel restrictions could delay the arrival and spread of epidemics and that select isolated locations may benefit from border closures. Once again, however, the overall size of the effects was relatively small and the quality of evidence was found to be very low.^10^ Another review of both international and domestic travel restrictions concluded that such measures could delay, but not contain, dissemination of both pandemic and seasonal influenza after it emerged.^11^ Based on the 23 studies identified, the review concluded that internal and international border restrictions could delay the spread of an outbreak by one week and two months, respectively, and that such restrictions could delay the spread and peak of epidemics from between a few days to up to four months. However, the timing of the introduction of such measures was key – the extent of the delay of spread was greatly reduced when restrictions were imposed more than six weeks after the onset of an epidemic.

Beyond influenza, evidence from other infectious disease outbreaks is more limited. A modelling study of travel restrictions implemented during the West African Ebola outbreak estimated that they may have delayed further international transmission by a few weeks for some countries.^12^ Given the low proportion of all international travellers originating in Ebola-affected countries at that time, another study suggested that exit screening measures in affected countries were likely to be more effective at reducing onward international transmission than travel restrictions,^13^ a finding that was supported by another similar study.^14^ The travel advisories issued by WHO during the 2003 SARS outbreak, which led to substantial declines in international travel to Hong Kong and Mainland China, were estimated to have delayed the export of cases by only a few days.^15^ Importantly, other studies have suggested that travel measures during outbreaks can be counter-productive by preventing countries from launching effective epidemic responses,^16^ undermining the detection of cases, and causing widespread economic effects on the travel industry itself.^17^ Since the onset of the pandemic, it has become clear that the clinical features of COVID-19 make it more challenging than previous infectious diseases to detect and contain,^18^ raising questions about whether evidence of effectiveness from previous studies is even relevant for COVID-19.^19^

The goal of this paper is to review evidence of the effectiveness of travel-related measures implemented during the early stages of the COVID-19 pandemic, a time of many unknowns regarding the clinical and epidemiological features of the novel coronavirus. Since the emergence of COVID-19, dozens of studies have been published or made available that evaluate the effectiveness of travel-related measures in the context of the pandemic. A recent Cochrane review of the literature on the effectiveness of international travel-related measures to contain COVID-19, severe acute respiratory syndrome (SARS), and Middle-East respiratory syndrome (MERS) identified 36 unique studies, of which only 25 were specific to COVID-19. The review concluded that cross-border travel measures may limit the spread of disease across national borders, specifically in terms of reducing the number of imported cases and delaying or reducing epidemic development, although it found that the certainty of the reviewed evidence was low to very low. Importantly, it did not consider travel-related measures adopted within China at the outset of the pandemic or studies that evaluate interventions implemented in combination. Given the widespread adoption of travel restrictions, and the likely enormous economic and social consequences resulting from them, a fuller understanding of the effectiveness of all of the measures adopted during the early phase of the outbreak is warranted. While the question of whether the adoption of these measures is compliant with the IHR has received attention in the literature,^20,21^ it is beyond the scope of this paper.

## Methods

To conduct this review, we adopted an abbreviated version of the preferred reporting items for systematic reviews and meta-analysis protocol (PRISMA-P) using the 17-point checklist.^22^ The rationale for the study was the widespread adoption of travel-measures despite consensus view at the time that such measures were largely ineffective, and in order to strengthen the application of the IHR during this and future pandemics. The objectives were to rapidly review evidence of the effectiveness of the full range of travel-measures adopted during the early stage of the COVID-19 pandemic form both published and unpublished studies. We further elaborate on other methods of the study below.

### 2.1 Search Strategy

Our search strategy was designed to be as inclusive as possible of all studies (as of June 1, 2020) that provide new evidence of the effectiveness of any travel-related measure adopted during the early phase of the COVID-19 pandemic. According to the IHR, travel-related measures include travel advice, entry and exit screening of travellers, medical examination or vaccination requirements for travellers, isolation or quarantine of suspected or affected persons, the refusal of entry of travellers, and restrictions on travellers from affected areas.^1^ While only those measures that are applied by State Parties at the level of an international border are covered by the IHR, many of these measures have also been applied to domestic travellers (e.g. at the level of inter-provincial or inter-state borders). Private companies, such as airlines and cruise ships, have also implemented travel measures, which, while also not subject to the IHR, further restricted travel during this pandemic. We did not restrict the search to specific outcomes (e.g. epidemiological or otherwise), or any specific methodological approach, or any specific geography. We only identified one study that looked at non-epidemiological outcomes,^23^ so while we include it in our description of the search and screening strategies, we exclude it from our main analysis below.

Given the rapidly evolving nature of the outbreak, as well as the rapidly expanding published literature on COVID-19, our search strategy targeted both pre-print and published articles, with the strategies to identify each differing slightly. Keywords were identified based on both inductive iterative testing of potential keywords, as well as deductively through papers identified through other channels. Search terms were then refined to minimize overlap and to maximize the number of studies that could be identified. While we did not impose a language restriction, we did not specifically search in non-English sources.

For pre-print papers, we searched the BioRxiv and MedRxiv servers, which offer limited search functionality, using the following keywords in the title field: travel*, flight*, airline*, border*, airport*, passenger or air traffic. We restricted the sample to papers that also included at least one COVID-19 keyword, either related to the virus itself (e.g. coronavirus, corona virus, coronavirinae, coronaviridae, betacoronavirus, covid19, covid 19, covid-19, nCoV, CoV 2, CoV2, sarscov2, 2019nCoV, novel CoV, OR Wuhan virus), the location of the early outbreak (e.g. Wuhan, Hubei or Hunan) or less specific but widely used terms (severe acute respiratory OR pneumonia AND outbreak). We used the same travel-related keywords to search the WHO’s COVID-19 global research database but did not impose a COVID-19 search term as theoretically all articles in this database were on this topic.^b^

For published papers, we searched PubMed with the following search strategy: studies must include at least one COVID-19 keyword mentioned above or one of the location-specific terms mentioned above combined with either (“severe acute respiratory” OR pneumonia AND outbreak) or one of the following MESH terms (Coronavirus, Coronavirus Infections, or Betacoronavirus) or the Supplementary Concept (COVID-19 or severe acute respiratory syndrome coronavirus 2). All studies must in addition include at least one travel-related measure term in the title or abstract (e.g. screening, travel advice, travel advisory, cordon sanitaire, ban, restrict*, prohib*, or clos*) as well as one travel-related term in the title or abstract (travel*, cruis*, ship*, terrest*, airplane, flight, plane, migrant, passenger, return, outflow, outbound, inbound, inflow, traffic, arrival, train, trains, bus, buses, transit, port*, airport*, tourist*, international importation, international exportation, case importation, imported cases, exported cases, or border).

### 2.2. Screening strategy

As of June 1, 2020, we identified a total of 312 articles, and during the review process, we identified another 8 articles through other sources (see Figure 1). From all identified studies (n=320), we removed duplicates, which left 300 articles to be screened. We uploaded the abstracts into Covidence^c^, software developed for systematic reviews, to perform a title and abstract screen. Our inclusion criteria were that studies must investigate the COVID-19 pandemic and at least one travel-related measure (applied either at an international or domestic border), must be empirical (i.e. modelled or observational), and must evaluate a specific outcome (epidemiological or other). Measures could have been undertaken by either a public (i.e. a government) or a private actor (e.g. an airline). We excluded articles that were news reports, review articles, commentaries or editorials, or conjecture (i.e. did not provide new data or evidence) about the effectiveness of travel measures. Each article was screened by 2 independent reviewers (TLH, LZH). Where there was disagreement among the reviewers, a third reviewer (KAG) resolved any disagreement.

**Figure 1:**
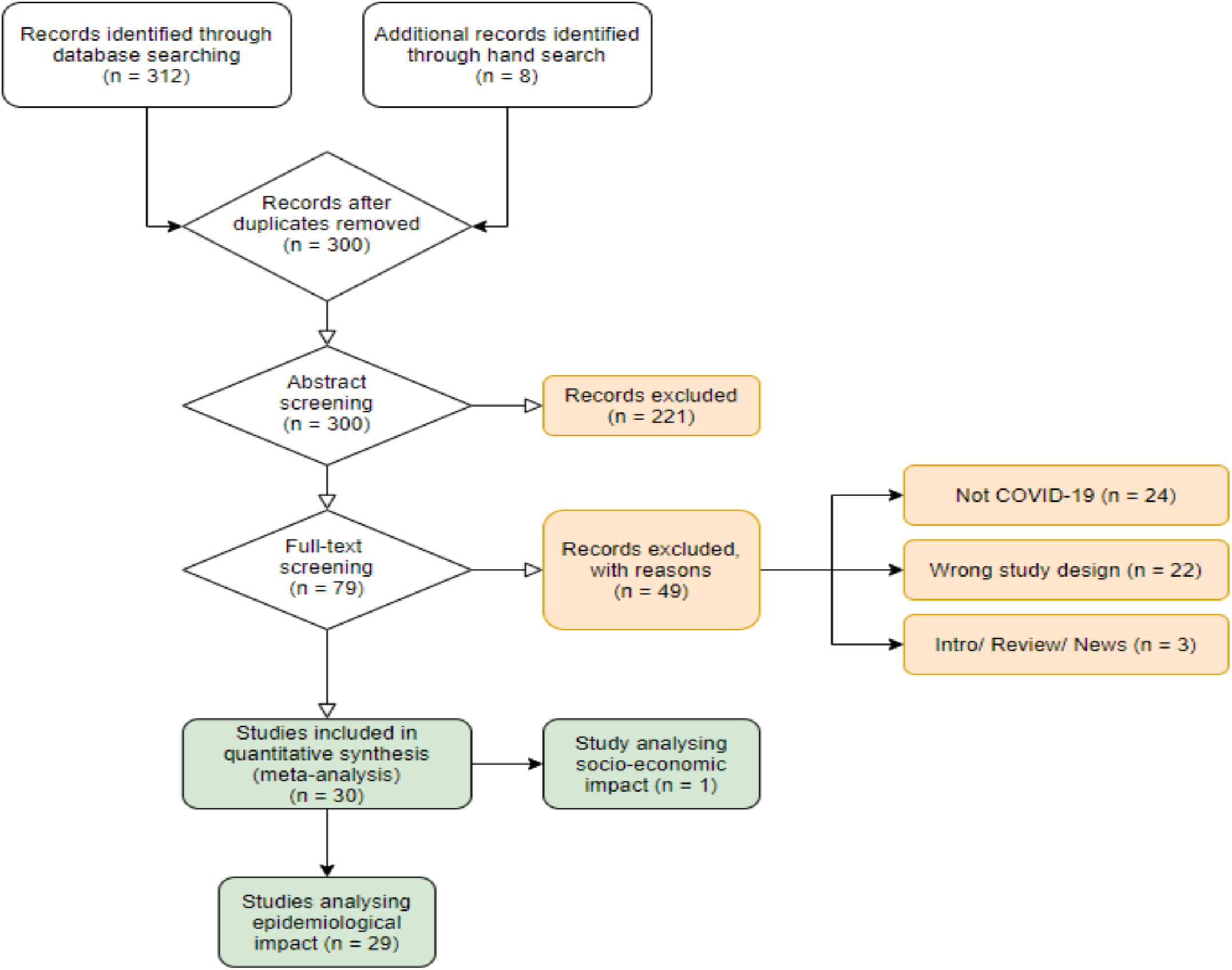
Prisma Diagram

After screening, we were left with 79 articles. The full texts of each were obtained and again uploaded into Covidence. Two reviewers again screened each article to determine if the article still met the inclusion criteria. Any disagreements by the two reviewers were again by a third reviewer (KAG) who was not otherwise involved in the screening process. After the full-text screening, we were left with 30 studies that met all of our selection criteria. Of these, one study investigated socio-economic outcomes, which we exclude from this analysis given the lack of overlap in outcomes.

### 2.3 Updating strategy

We continued to search the relevant databases for any newly published papers or to identify papers that had originally been identified as pre-prints but were subsequently published, until 1 June 2020. We retained the subsequently published versions of such papers.

### 2.4 Data extraction

For the remaining 29 studies, we extracted the title, authors, article source, publication date, whether it was a pre-print or published (or had previously been a pre-print) article, country context in which the study was conducted (or global), country(ies) implementing the measure, the country(ies) affected by the travel measure, specific measure(s) adopted, the timing of the measures, the duration of the measures, whether the study was modeled or based on observational data, the type of model used, epidemiological assumptions made in the models, the specific outcomes observed, the overall findings, the way in which cases/deaths were recorded, whether there was any description of diagnostic methods used to identify cases/deaths, whether the study made assumptions about asymptomatic cases, whether the study also accounted for secondary transmission, the extent to which the model also accounted for other measures imposed around the same time, the data sources used in the study, and the stated limitations of each study. For modelled studies, we also collected the name of the model used if an existing model was used, whether the model used was a dynamic or static model, whether the model used was a stochastic or deterministic model, and whether it was an individual vs. population-based model. A full list of the papers is presented in Appendix Table 2.

**Table 1:** Summary Statistics.

|  | Pre-Print | Published | Total |
| --- | --- | --- | --- |
| **Date of pre-print submission/ publication** |  |  |  |
| January | 0 | 1 | 1 |
| February | 0 | 4 | 4 |
| March | 4 | 11 | 15 |
| April | 0 | 3 | 3 |
| May | 1 | 5 | 6 |
| **Study design** |  |  |  |
| Modelled | 5 | 21 | 26 |
| Observational | 0 | 3 | 3 |
| **Level of region affected by travel measures** |  |  |  |
| Mainland China | 3 | 14 | 17 |
| Other single country | 1 | 4 | 5 |
| Multi-countries or global | 1 | 6 | 7 |
| **Level of travel measures imposed** |  |  |  |
| International | 2 | 10 | 12 |
| Interprovincial | 3 | 13 | 16 |
| Both<sup>a</sup> | 0 | 1 | 1 |
| **Travel measures analysed<sup>b</sup>** |  |  |  |
| Suspended transportation | 4 | 20 | 24 |
| Border restrictions | 3 | 18 | 21 |
| Screening | 0 | 5 | 5 |
| Entry quarantine | 1 | 3 | 4 |

Note:
<sup>a</sup> papers evaluating the impact of both international and interprovincial measures.
<sup>b</sup> papers may analyse effects of multiple restrictions.

**Table 2:**
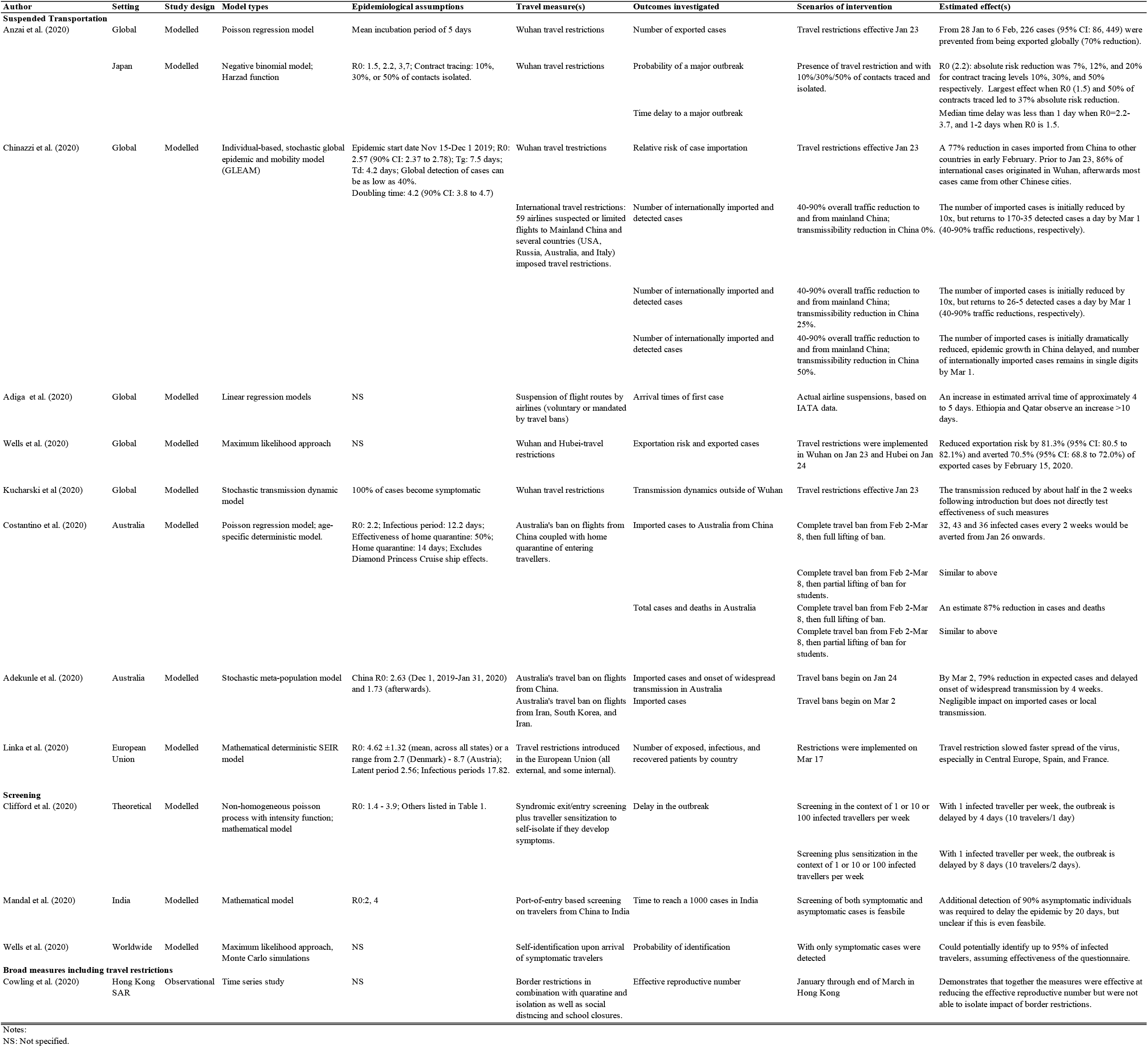
Effect of International Travel Measures.

### 2.5. Analysis of travel measures

We characterized the investigated travel-related measures into four groups: suspended transportation, border restrictions, entry or exit screening, and entry quarantine, which are summarized in Table 1. Papers may have investigated more than one measure and thus may contribute more than once to the table. In our analysis, we grouped studies according to whether the measure affected international (Table 2) or domestic travellers (Table 3). One study fit both criteria and thus is listed twice in the analysis. We used a narrative approach to synthesize the evidence of effectiveness. Two studies,^24,25^ despite study design and state objectives to investigate the impact of travel related measures, did not present their findings in such away that allowed us to extract the evidence generated in the study. These are summarized in appendix table 2 but not in tables 2 or 3.

**Table 3:** Effect of Implementation of Interprovincial Travel Measures.

| Author | Study design | Model type | Epidemiological assumptions | Travel measure(s) | Outcomes investigated | Scenarios of Intervention | Estimated effect(s) |
| --- | --- | --- | --- | --- | --- | --- | --- |
| Aleta et al. (2020) | Modelled | Stochastic SEIR-metapopulation model | Generation time: 7.5 days; $R_0$: 2.4; later period: 3 days | Wuhan travel measures | Cases in Mainland China outside of Wuhan | Travel measures implemented on Jan 23 | A reduced reduced number of cases but only in the short term. |
| Chinazzi et al. (2020) | Modelled | Individual-based, stochastic global epidemic and mobility model (GLEAM) | $R_0$: 2.57; doubling time: 4.2; no changes in transmissibility within China | Wuhan travel measures | Cases in Mainland China outside of Wuhan<br>Timing of epidemic peak | Travel measures implemented on Jan 23 | Reduction of cases was approximately 10% by 31 January (range 1 to 58%).<br>Wuhan travel ban delayed epidemic progression by 3 to 5 days in China. |
| Fang et al. (2020) | Modelled | Dynamic distributed lag regression model | Incubation period: up to 22 days | Wuhan travel measures | Number of cases in cities outside of Hubei from Jan 23-Feb 29 | Travel measures implemented on Jan 23 | COVID-19 cases would be 64.81% higher in 347 cities outside Hubei (20,810 v. 12,626), and 52.64% higher in 16 other cities in Hubei as of 29 February (23,400 v. 15,330). |
| Shi et al. (2020) | Modelled | Autoregressive integrated moving average (ARIMA) model | Incubation period: 4-6 days | Wuhan travel measures | Cumulative number of confirmed cases outside Wuhan by Feb 29 | Travel measures implemented on Jan 23 | Travel ban may have prevented approximately 19,768 (95% CI: 13589, 25946) cases outside of Wuhan by Feb 29 (39% reduction). |
| Tian et al. (2020) | Modelled | Deterministic SEIR model | $R_0$: 3.15 | Wuhan travel measures | Arrival times in cities across Mainland China by Feb 19<br>Cases in Mainland China outside of Wuhan by Feb 19 | Travel measures implemented on Jan 23 | Delayed arrival time by 2.91 days (95% CI: 2.54, 3.29).<br>National number of cases decreased from 744,000 (±156,000) to 202,000 (±10,000) (72.8% decrease). |
| Kraemer et al. (2020) | Modelled | Generalized linear model | Doubling time: 4.0 days (outside Hubei), 7.2 days (inside Hubei); incubation period 5.1 days | Wuhan travel measures | Cases in Mainland China outside of Wuhan by end of Feb | Travel measures implemented on Jan 23 | Travel measures reduced growth rates outside, which became negative after Jan 23; Provinces with greater mobility from Wuhan displayed more rapidly declining growth rates. |
| Tang et al. (2020) | Modelled | Deterministic SEIR model | $R_0$: 6.47 | Wuhan travel measures | Cases in select locations outside of Wuhan | Travel measures implemented on Jan 23 | Travel restriction reduced the number of infected individuals in Beijing over 7 days by 91.14%. |
| Hou et al. (2020) | Modelled | Deterministic SEIR model | Incubation period: 7 days | Wuhan travel measures | Effective reproductive rate | Travel measures implemented on Jan 23 | Travel measures significantly changed transmission dynamics within China |
| Li et al. (2020) | Modelled | Stochastic SEIR model | $R_0$ at the beginning of the epidemic to be 2.38 | Wuhan travel measures | Reproductive number | Travel measures implemented on Jan 23 | Travel measure reduced the reproductive number from 2.38 down to 1.34 and 0.98, in the 1 and 2 week period immediately following their introduction. |
| Lau et al. (2020) | Observational | Retrospective regression model | $R_0$: 2.2-3.9; mean incubation period: 5.1 days | Wuhan travel measures | Doubling time | Travel measures implemented on Jan 23 | Significant increase in doubling time from 2 to 4 days after lockdown. |
| Wu et al. (2020) | Modelled | Deterministic SEIR model | $R_0$=2.6, zoonotic force=86/day until Jan 1 market closure | Wuhan travel measures | Exported cases in the rest of Mainland China | Travel restrictions led to either a 0% or 50% reduction in travel outside of Wuhan | Even a 50% reduction in inter-city mobility would have a negligible effect on the epidemic dynamics. |
| Liu et al. (2020) | Observational | Linear regression | Incubation period: 3-7 days | Wuhan travel measures | Cases exported outside of Wuhan | Travel measures implemented on Jan 23<br>Travel measures implemented 2 days earlier<br>Travel measures implemented 2 days later | A mean values of 129 cases exported per 10,000 people who left Wuhan<br>An estimated 1420 (95% CI: 1059, 1833) cases would have been prevented.<br>An estimated 1462 (95% CI: 1059, 1833) additional cases would have happened. |
| Yuan et al. (2020) | Modelled | Regression model | Incubation period: 5 days | Wuhan travel measures in combination with a stay at home movement | Cases in 44 regions outside of Wuhan by Feb 27<br>Cases in 44 regions outside of Wuhan by Feb 27 | Travel measures implemented on Jan 23<br>Travel measures implemented three days earlier | Reduced the number of cases outside of Wuhan from 41,477 to 30,765.<br>Further reduce the number of cases to between 15,768-21,245. |
| Su et al. (2020) | Modelled | Deterministic SEIR model | $R_0$ = 2.91, 2.78, 2.02, and 1.75 for Beijing, Shanghai, Guangzhou, and Shenzhen respectively | Wuhan travel measures in combination with other non-pharmaceutical interventions | Transmission rates in four metropolitan areas of China | Different contract rates were assumed to result from reduced population flow. | Travel restrictions contributed to a reduction in the contact rate and reduced the time to peak and the number of cases. |
| Jiang et al. (2020) | Modelled | Time-varying sparse vector autoregressive model | Incubation period: 10 days | Travel measures introduced in 5 cities of Hubei (Wuhan, Huanggang, Ezhou, Chibi and Zhijiang) | Daily transmission routes from Hubei to other provinces through Feb 19 | Travel measures started to be implemented on Jan 23 | Travel restrictions reduced transmission between provinces |
| Lai et al. (2020) | Modelled | Stochastic SEIR model | $R_0$: 2.2; incubation period: 5.2 days | Wuhan travel measures in combination with other non-pharmaceutical interventions | Cases in Mainland China outside of Wuhan<br>Cases in Mainland China outside of Wuhan<br>Total number of cases outside of Wuhan | Travel measures implemented on Jan 23<br>If travel restrictions of same magnitude were implemented one, two, or three weeks earlier<br>If travel restrictions of same magnitude were implemented one, two, or three weeks later. | Early detection and isolation of cases more effective than travel restrictions; travel restricts reduced the number of cases outside of Wuhan as well as its geographic spread.<br>If interventions were conducted one, two, or three weeks earlier, cases will reduce by 66%, 86%, or 95% respectively.<br>If interventions were conducted one, two, or three weeks later, cases may increase 3-fold, 7-fold, or 18-fold respectively |
| Hossain et al. (2020) | Modelled | Meta-population model | $R_0$: 2.92; latent period: 5.2 days; generation time: 8.4 days | Border control and quarantine | Arrival time outside of Wuhan | Theoretical application of measures | Arrival time is delayed by 32.5 days and 44 days under a low $R_0$ (1.4) but under higher $R_0$ (2.92) only 10 extra days can be gained. |

### 2.6. Analysis of outcome measures

Outcomes included the number of observed cases, date of the epidemic peak, risk of transmission, case growth rate, doubling time, time of arrival in a new country, the reproductive number (R_0_ or R_t_), and projected cumulative cases. Details are listed in Table 2 and Table 3, which summarize papers that investigate international and domestic travel measures respectively. One article that evaluated the effectiveness of both types of measures,^26^ appears in both tables.

### 2.7 Assessment of bias

Individual articles were assessed for bias using a proprietary scoring system consisting of three tiers: low, moderate, and high risk of bias. Low scores were given when the author had adequately addressed the domain, moderate when it had been either partially or incompletely addressed, or high when it was not or only poorly addressed. Articles were assessed with regards to their ability to form a clear and precise definition of the research question, travel restriction measures included in the analysis, comprehensiveness of outcome, suitable mathematical modelling, model assumptions described, confounding factors, model validation, and uncertainty assessment.^27^ A detailed summary of our bias assessment is presented in Appendix Table 1.

## Results

Of the 29 identified studies in this rapid review, 24 had been published by September 1, 2020, while the rest were pre-print studies (see Table 1). Almost all of the studies (26) were modelled studies with few observational studies (3). Given the timing of the studies, almost all of the studies focused on the initial exportation of cases from Wuhan either domestically within China or internationally. Among the travel-related measures adopted, the most commonly investigated were suspended transportation (24), border restrictions (21), and screening (5). Only four studies investigated entry quarantine.

### Effectiveness of travel-related measures on the international spread of COVID-19

In Table 2, we present the summary of evidence generated from the papers that investigated the impact of international travel measures. All but one^28^ were modelled studies. Four studies directly investigated the impact of the Wuhan travel ban on the initial export of cases internationally.^26,29–31^ Comparing the observed number of exported cases to scenario-based modelled estimates without the ban, studies consistently found that these measures were highly effective at reducing exportation of cases. Anzai et al. estimated a 70% reduction in exported cases globally in the week following the introduction of the ban,^29^ Chinazzi et al. estimated that the ban led to a 77% reduction in imported cases,^26^ while Wells et al. estimated that it reduced the risk of exportation by about 80% through mid-February.^30^ Kucharski estimated that transmission rates outside of China were reduced by about half in the two weeks following the introduction of the ban.^31^ However, Anzai et al., which focused on the impact of the ban on the outbreak in Japan, estimated that the absolute risk of a major outbreak was only modestly delayed due to the Wuhan travel ban and that the median time delay in a major outbreak was only 1-2 days.^29^

Beyond the direct effect of the Wuhan travel ban, Chinazzi et al. estimated that the application of additional travel-related restrictions, on travellers to and from Mainland China by receiving countries, led to additional reductions in imported cases globally, though the extent of reduction varied by country.^26^ Adiga et al. investigated the impact of government or airline-imposed travel-related measures and estimated that these led to a delay in the importation of the virus by about 4-5 days on average and up to 10 days in select countries.^32^ This study, however, did not directly control for the impact of the Wuhan lockdown, which happened around the same time as many of the measures investigated.

In terms of specific country case studies, Adekunle et al. found that Australia’s ban on air travel to and from China may have prevented 82% of imported cases through February 2.^33^ Similarly, Costantino et al. estimated that it may have led to a 79% reduction in imported cases through March 2.^34^ Linka et al. estimated that the travel restrictions implemented both at the external and internal borders of the European Union significantly decreased the speed of virus spread across member states, especially in Central European countries.^35^ These last two studies, both of which focused on an earlier period in the pandemic, failed to account for the impact of the Wuhan lockdown in addition to the restrictions evaluated.

Studies that investigated the effectiveness of screening found that only very highly effective screening could reduce [or decrease] the risk of importation or exportation. Clifford et al. found that when the number of cases was low in the exporting country, screening may delay the onset of the epidemic in the importing country by up to a week^36^ while Mandal et al. found that if screening could detect 90% of asymptomatic individuals, it could delay the average time of the epidemic by up to 20 days in select countries.^37^Assuming that self-identification of cases was effective, another study suggested that such measures could identify a large proportion of infected travellers. However, this assumes that screening is effective but does not study this directly.^30^

A single observational study identified in this review investigated the impact of border restrictions, in combination with mandatory quarantine and screening, for incoming travellers to Hong Kong. Cowling et al. concluded that the application of quarantine measures of incoming travellers into the region was an important element of their successful public health response, but the study does not specifically estimate its impact independent of other measures including travel-related measures.^28^

### Effectiveness of travel-related measures on the domestic spread of COVID-19 within China

In Table 3, we present the findings from the studies that investigated the impact of the Wuhan travel restrictions, on the domestic export of cases to other parts of China. By comparing actual observed cases to counterfactual scenarios where such measures had not been imposed, Chinazzi et al. predicted that the travel ban led to a 10% reduction of exported cases within the first 7 days,^38^ Fang et al. estimated a 39.3% reduction over one month,^38^ Shi et al. similarly identified a 39% reduction in cases over one month,^39^ while Tian et al. estimated a 73% reduction through mid-February.^40^ Tang et al. found that the Wuhan travel ban led to a 91.1% reduction in imported cases in Beijing over 7 days.^41^ Similarly, Kraemer et al. also found that these travel measures dramatically reduced the transmission of the outbreak across the country, with areas which had greater pre-lockdown connectivity with Wuhan experiencing a greater decline.^42^ Aleta et al. estimated that the measures were effective in reducing the exportation of cases, but only in the short term.^43^ Yuan also found the lockdown to be effective at reducing the number of cases outside of Wuhan but notes that the timing also coincided with a nationwide stay at home campaign imposed by the central Chinese government.^44^

Studies also investigated the impact of the travel ban on the domestic timing of the outbreak. Tian at el. estimated the ban delayed outbreaks within China by 2.91 days,^40^ while Chinazzi et al. estimated a delay of 3 to 5 days.^26^ Studies also investigated the impact of the travel ban on the effective reproductive rate, the doubling time, and other measures. Hou et al. found that the ban quickly reduced the reproductive rate of the virus outside of Wuhan,^45^ similarly Li et al. found that the reproductive number dropped by more than half within 2 weeks of the introduction of the ban,^46^ Using observational data, Lau et al found that the doubling time of the virus increased from 2 days to 4 days after the travel ban was imposed.^47^ Another study found substantial declines in transmission routes between Chinese provinces within weeks of the introduction of the Hubei travel bans.

The timing of travel-related measures, again, appears to be important in predicting effectiveness. Both Lai et al. and Liu et al., the former a modelled study and the latter an observational study, estimated that the Wuhan travel ban would have been substantially more effective if implemented 1-3 weeks earlier.^48,49^ This is supported by Wu et al. who found that the travel ban had a relatively minor effect on the overall speed of transmission of the outbreak in areas of China outside of Wuhan, largely because a large number of cases had already been exported before the travel ban, limiting its effectiveness.^50^

Finally, a few studies evaluated domestic travel bans in combination with other travel-related measures. For example, one modelled study estimated that if major cities within China had imposed additional measures, they could have further reduced their epidemic risk.^51^

### Quality of available evidence

Our review of the risk of bias in the included studies (Appendix Table 1) suggests that, apart from a few exceptions, while most of the studies had clear research questions, descriptions of the travel-related measure(s) evaluated, and discussions of the outcomes, few of the studies made efforts to adequately control for the presence of other public health or travel-related measures implemented at the same time, or for other contextual factors that could influence the impact of travel-related measures. One important challenge common to all of the studies was the quality of data on detected cases early on in the pandemic and only a small number of the studies allowed for their estimates to vary based on potential ranges of the number of true cases that were actually detected early in the outbreak (e.g. Chinazzi et al. estimated 24.4% of all cases were undetected,^26^ Fang et al. estimated 42.0-80.0% were undetected,^38^ Kucharski et al. predicted that there were at least ten times as many cases as were confirmed in Wuhan in early February^31^). The studies also varied markedly in their efforts to validate their models or provide uncertainty analysis around their estimates. Also, with the exception of a few studies, most studies did not discuss the potential measurement error associated with case data collected during the early phases of the pandemic which likely did not capture most asymptomatic cases. The overall quality of the studies to evaluate effectiveness was thus relatively low.

## Discussion

Despite WHO’s recommendations against travel restrictions, and given limited evidence of the effectiveness of such measures at the onset of the COVID-19 pandemic, there has been an unprecedented adoption of such measures, both domestically and internationally, which has led to dramatic declines in international travel. This paper reviews the emergent evidence on the effectiveness of travel-measures adopted during the early phase of the COVID-19 pandemic.

A number of key findings emerge from this rapid review of the effectiveness of measured adopted in the early phase of the pandemic. First, there was a high level of agreement among the studies that the adoption of travel measures played an important role in shaping the early transmission dynamics of the COVID-19 pandemic. However, almost all the studies in this review focused on either domestic or international travel bans imposed on Wuhan, and to a lesser extent the rest of China, during the early period of the pandemic. This review does not identify substantial new evidence of the effectiveness of travel-related measures aimed at controlling spread to and from other parts of the world.

Second, the evidence suggests that the Wuhan travel measures were likely effective at reducing the initial exportation of the virus within China and abroad. At the international level, studies consistently estimated that these measures led to a 70-80% reduction in cases exported in the first few weeks, and likely had a smaller effect within Mainland China, where estimates of effectiveness ranged from 10-70%. Also, the Wuhan travel ban likely led to delays of up to a few weeks in the importation of cases to other countries. However, most of the studies also concurred that the effects were short-lived. This suggests travel-related measures alone are unlikely to significantly change the trajectory of the outbreak unless commensurate domestic measures are also implemented. This is supported by evidence that once four or more infections are introduced into a new location, there is an over 50% chance that a major outbreak will occur.^31^

Third, the studies reviewed suggest a key factor influencing the effectiveness of travel-related measures was timing. Within China, had the same policies been implemented a few weeks earlier, it is likely there may have been substantially less seeding of new outbreaks across the country and internationally. While the measures aimed at Wuhan significantly reduced the exportation of cases from the province, over time other provinces became the source of most of the internationally exported cases. Where authorities are not able or willing to adopt such measures at a sufficiently early stage of an outbreak, once again, this suggests the need to give commensurate attention to strengthening the role of other domestic measures such as testing, contact tracing, and physical distancing.

Fourth, many of the studies focused on international travel failed to account for the implementation of the domestic Wuhan travel ban. Although this was a domestic policy, and thus outside the remit of the IHR, evidence suggests restricting travel to and from Wuhan dramatically changed the outflow of cases from the region at a crucial time period. Studies that did not account for the Wuhan travel ban in their estimates, or reviews that excluded domestic travel measures, thus likely overestimated the effectiveness of international travel measures. The notable exception was Chinazzi et al. who conclude that the additional measures provided a benefit above and beyond the effect of the Wuhan measures, at least in the short run.^26^

Fifth, during the early phase of the pandemic there were likely large numbers of undetected cases globally, and although some studies allowed for their estimates to vary based on assumed proportions of undetected cases, the validity of the estimated effects in all of these studies is likely affected by data quality issues. Also, given that symptomatic individuals may be more likely to curb their travel than asymptomatic travellers, especially internationally and after the introduction of travel-related measures aimed at detecting symptomatic cases, modelled effectiveness studies may also be biased if they do not account for this difference. In addition, it is also possible that some of the travel-related measures adopted (e.g. screening) could have actually led to increased detection of cases,^46^ which could further complicate the ability of studies to evaluate effectiveness as the measurement of the outcome is influenced by the intervention itself and few studies acknowledged this limitation.

Finally, while this study identified a relatively large number of studies, we assess the quality of these studies overall to be low. Almost all of the studies identified in this review were modelled studies, and therefore the results depend upon important parameter assumptions, which varied considerably. Given the rapidly evolving and dynamic nature of the pandemic, it is unclear how close to reality these assumptions were. Comparability across the studies is also undermined by a lack of standardized terminology. Furthermore, few studies attempted to isolate the potential effect of international travel-related measures from a range of domestic measures implemented concurrently, as well as from other social, political, or economic characteristics of the implementing or target locations or populations.

This systematic review also has several important limitations of its own. First, while we aimed to be systematic in our search strategy as well as inclusion criteria, the rapidly expanding literature on COVID-19 pandemic almost certainly means that we likely overlooked some relevant studies. Second, although we aimed to focus on the early phase of the outbreak, it is unclear when the appropriate time was to end our review. Newer studies that have been published since we ended searching the literature may present a different picture on the effectiveness of travel measures and thus the evidence from this study must be evaluated in this context. Third, assessments of bias in studies is challenging and is inherently subjective in nature.

In recent months, a process has commenced to review and strengthen the IHR. The universal use of travel-related measures by States Parties during the COVID-19 pandemic will be a major focus of discussion on the limitations of the current treaty. Based on this review, we make the following recommendations. First, the findings of this review suggest that it is difficult to know in the early phases of an infectious disease outbreak how effective travel measures are likely to be, based on evidence from previous studies. Overall, the findings of the review suggest that while it is likely that travel measures can affect the early dynamics of an infectious disease outbreak, their effectiveness in delaying or reducing international spread is likely to be limited on their own and short-lived. But the review also finds that the way in which these measures are adopted likely influences their effectiveness. This highlights the need to from blanket assessments of the effectiveness of travel-related measures (“travel measures don’t work”), to context-specific assessments of effectiveness based on scenarios of transmission risk (“when might such measures be effective”?).

Second, it is also clear that the effectiveness of travel-measures cannot be estimated using a single fixed parameter. The effectiveness of measures will vary based on the setting, which other measures are also implemented, the extent to which they are implemented, and the speed at which they are implemented. All of these factors, weighed against potential harms, need to be taken into consideration in discussions about the potential effectiveness of international travel measures.

Third, this study finds that measures implemented early were likely more effective than those implemented late. In this pandemic, the WHO did not make any recommendations on travel restrictions until it had issued the PHEIC, weeks after when many countries had already implemented such measures, and when it did, it had recommended against such measures based on the limited evidence available at the time. In addition, the IHR require State Parties to provide evidence for any additional health measures that they implement. In the context of an outbreak of a novel infectious disease agent, it is unclear what justifies evidence in the early phases of the outbreak. The role of evidence in the IHR in such settings therefore needs to be reconsidered.

This review also highlights areas where more research is urgently needed to understand the appropriate role of travel measures during PHEICs. First, a greater understanding of effectiveness of travel measures adopted globally, and at stages beyond the early phases of the pandemic are urgently needed. Second, there is also a need to better understand the broad range of measures affecting travel and trade beyond those covered in the reviewed studies, including the role of testing, which was not widely available during the early phase of the pandemic. Third, lack of data on true case numbers remains an underlying challenge across all of the study reviewed, and thus needs to be taken more seriously in future studies. Fourth, models need to better account for the way in which travel measures work in tandem with other measures implemented concurrently, including domestic travel measures and other public health measures. Fifth, studies need to better account for the possibility that some locations see grater benefit from travel measures than others based on geographic and socioeconomic factors. Finally, one of the rationales against the use of travel measures is their economic and social impacts, yet few studies in the early phases evaluated their non-epidemiological outcomes.

At the onset of the COVID-19 pandemic, there was a widely held belief that travel measures were unlikely to play much of a role in curbing international spread of the virus. However, the widespread adoption, and persistent use of such measures, globally, as well as some of the evidence identified in this review, challenge this belief. While this review emphasizes that the quality of evidence remains low and highlights a number of methodological shortcomings in the reviewed studies, this review also identified new evidence of the impact of such measures during the early phase of the pandemic. These findings suggest such measures did play an important role in shaping early dynamics of the pandemic, even if they were unable to contain the virus globally on their own. Like with so many things that COVID-19 has transformed, and the pandemic has also challenged our views of what constitutes evidence of effectiveness of public health measures.^52^

## Supporting information

PRISM-P checklist

## Data Availability

We used publically available data.

## Role of the Funding Agency

The authors are funded by the New Frontiers in Research Fund (Grant NFRFR-2019-00009) through an operating grant awarded under the Canadian Institutes of Health Research Rapid Research Funding Opportunity. The funders were not involved in the design or writing of this study.

## Contributions

KG developed the study methodology, drafted the manuscript, and oversaw data analysis. ZL and TLH conducted the systematic review search, article screening, data extraction, and data analysis. KL, CZW, and SM contributed to the design of the study and provided input into the manuscript.

---

**Appendix Table 1:** Risk of Bias Assessment.

| Authors | Clear and precise definition of research question | Travel restriction measures included in analysis | Outcomes | Suitable mathematical modelling | Major model assumptions described | Other related factors adjusted | Model validated | Uncertainty analysis |
| --- | --- | --- | --- | --- | --- | --- | --- | --- |
| Anzai et al. (2020) | Low | Low | Low | Low | Low | Moderate | High | High |
| Adekunle et al. (2020) | Low | Low | Low | Moderate | Low | Moderate | Low | Low |
| Adiga et al. (2020) | Low | Moderate | Low | Low | Low | High | Low | High |
| Aleta et al. (2020) | Low | Low | Low | Moderate | Low | High | Low | Low |
| Boldog et al. (2020) | Low | Low | Low | Low | Low | Moderate | High | Low |
| Chinazzi et al. (2020) | Low | Low | Low | Low | Low | Low | Low | Low |
| Clifford et al. (2020) | Low | Low | Low | Low | Low | High | High | Low |
| Costantino et al. (2020) | Low | Low | Low | Low | Low | Moderate | High | Moderate |
| Cowling et al. (2020) | Low | High | Low | Low | NA | High | NA | NA |
| Fang et al. (2020) | Low | Moderate | Low | Low | Low | Low | Low | Low |
| Hossain et al. (2020) | Low | Low | Low | Low | Low | Moderate | Low | High |
| Hou et al. (2020) | Low | High | Low | Low | Low | High | Low | Low |
| Jiang et al. (2020) | Low | High | Low | Low | Low | High | Low | Low |
| Klausner et al. (2020) | Low | High | High | High | High | High | High | High |
| Kraemer et al. (2020) | Low | Low | Low | Low | Low | Low | Low | Low |
| Kucharski et al. (2020) | Low | High | Low | Low | Low | High | Low | Low |
| Lai et al. (2020) | Low | Low | Low | Low | Low | Low | Low | Low |
| Lau et al. (2020) | Low | Moderate | Low | Low | Low | High | NA | NA |
| Li et al. (2020) | Low | High | Low | Low | Low | High | Low | Low |
| Linka et al. (2020) | Low | Low | Low | Low | Low | High | High | Low |
| Liu et al. (2020) | Low | High | Low | Low | Low | High | Low | Low |
| Mandal et al. (2020) | Low | Low | Low | Low | Low | Moderate | High | Low |
| Shi et al. (2020) | Low | High | Low | Low | Low | High | Low | Low |
| Su et al. (2020) | Low | High | Low | Low | Low | High | Low | High |
| Tang et al. (2020) | Low | Low | Low | Low | Low | High | Low | Low |
| Tian et al. (2020) | Low | Low | Low | Low | Low | Low | Low | Low |
| Wells et al. (2020) | Low | Low | Low | Low | Low | Low | Low | Low |
| Wu et al. (2020) | Low | Low | Low | Low | Low | Moderate | Low | Low |
| Yuan et al. (2020) | Low | Moderate | Low | Moderate | Low | High | Low | High |

**Appendix Table 2:** Summary of included papers.

| Author | Published/Pre-printed |  | International / Interprovincial | Geographical context | Location(s) implementing measure(s) | Traveler(s) affected by measures | Travel Measures |
| --- | --- | --- | --- | --- | --- | --- | --- |
| Anzai et al. (2020) | Published | Modelled | International | Global | Wuhan | Travelers from China and specifically travelers to Japan | Suspended air transportation |
| Adekunle et al. (2020) | Published | Modelled | International | Australia | Australia | Travelers from China, Iran, Italy, and South Korea to Australia | Suspended air transportation; Provincial border closure |
| Adiga et al. (2020) | Pre-printed | Modelled | International | Global | Airlines in 24 countries | Outbound travelers from China | Suspended air transportation |
| Aleta et al. (2020) | Published | Modelled | Interprovincial | Mainland China | Wuhan | Travelers within Mainland China | Suspended land, air, sea transportation; Provincial border closure |
| Boldog et al. (2020) | Published | Modelled | International | Global | Wuhan and other countries | Outbound travelers from China | Suspended land, air, sea transportation; Entry screening |
| Chinazzi et al. (2020) | Published | Modelled | Both | Global | Wuhan and other countries | Outbound travelers from China | Suspended air transportation; Provincial border closure |
| Clifford et al. (2020) | Published | Modelled | International | Global | Destination countries | Inbound travelers | Entry and exit screening |
| Costantino et al. (2020) | Published | Modelled | International | Australia | Australia | Travellers from China to Australia | Suspended air transportation |
| Cowling et al. (2020) | Published | Observational | International* | Hong Kong SAR | Hong Kong SAR | Inbound travelers | Suspended land, air, sea transportation; Entry screening; Entry quarantine |
| Fang et al. (2020) | Published | Modelled | Interprovincial | Mainland China | Wuhan | Travelers to 363 cities in China from Wuhan | Provincial border closure |
| Hossain et al. (2020) | Published | Modelled | Interprovincial | Mainland China | Wuhan | Travelers to major cities in China from Wuhan | Provincial border closure; Entry quarantine |
| Hou et al. (2020) | Pre-printed | Modelled | Interprovincial | Mainland China | Wuhan | Travelers within Mainland China | Suspended land, air, sea transportation; Provincial border closure |
| Jiang et al. (2020) | Published | Modelled | Interprovincial | Mainland China | Wuhan | Travelers within Mainland China | Suspended land, air, sea transportation; Provincial border closure |
| Klausner et al. (2020) | Pre-printed | Modelled | International | Israel | Israel | Inbound travelers, especially those from China | Entry quarantine |
| Kraemer et al. (2020) | Published | Modelled | Interprovincial | Mainland China | Hubei and other provinces | Travelers within Mainland China | Suspended land, air, sea transportation; Provincial border closure |
| Kucharski et al. (2020) | Published | Modelled | International | Mainland China | Wuhan | Outbound international travelers from Wuhan | Suspended land, air, sea transportation; Provincial border closure |
| Lai et al. (2020) | Published | Modelled | Interprovincial | Mainland China | Wuhan and other cities within China | Travelers within Mainland China | Suspended land, air, sea transportation; Provincial border closure |
| Lau et al. (2020) | Published | Observational | Interprovincial | Mainland China | Wuhan | Travelers within Mainland China | Suspended land, air, sea transportation; Provincial border closure |
| Li et al. (2020) | Published | Modelled | Interprovincial | Mainland China | Wuhan | Travelers within Mainland China | Suspended land, air, sea transportation; Provincial border closure |
| Linka et al. (2020) | Published | Modelled | International | Europe | European Union member countries | Travelers within the European Union | Suspended land, air, sea transportation; National border closure |
| Liu et al. (2020) | Published | Observational | Interprovincial | Mainland China | Wuhan | Travelers within Mainland China | Suspended land, air, sea transportation; National border closure |
| Mandal et al. (2020) | Published | Modelled | International | India | India | Inbound travelers to India | Entry screening |
| Shi et al. (2020) | Pre-printed | Modelled | Interprovincial | Mainland China | Wuhan | Travelers within Mainland China | Suspended land, air, sea transportation; Provincial border closure |
| Su et al. (2020) | Published | Modelled | Interprovincial | Mainland China | Provinces within China | Travelers within Mainland China | Suspended land, air, sea transportation; Provincial border closure |
| Tang et al. (2020) | Published | Modelled | Interprovincial | Mainland China | Wuhan | Travelers within Mainland China | Suspended land, air, sea transportation; Provincial border closure |
| Tian et al. (2020) | Published | Modelled | Interprovincial | Mainland China | Wuhan | Travelers within Mainland China | Suspended air transportation; Provincial border closure |
| Wells et al. (2020) | Published | Modelled | International | Global | Wuhan | Inbound international travelers from China | Suspended air transportation; Provincial border closure; Entry Screening; Entry Quarantine |
| Wu et al. (2020) | Published | Modelled | Interprovincial | Mainland China | Wuhan and Hubei province | Travelers within Mainland China | Suspended land, air, sea transportation; Provincial border closure |
| Yuan et al. (2020) | Pre-printed | Modelled | Interprovincial | Mainland China | Wuhan | Travelers within Mainland China | Suspended land, air, sea transportation; Provincial border closure |

\* Also includes measures to control border with Mainland China but does not distinguish between international and within China travelers so only listed in the international table.

## Footnotes

b https://www.who.int/emergencies/diseases/novel-coronavirus-2019/global-research-on-novel-coronavirus-2019-ncov

c https://www.covidence.org/

